# Protocol for a prospective cohort study on the feasibility of application of nutritional ultrasound in the diagnosis and follow-up of patients with nutritional risk at hospital discharge: Study on body composition and function (DRECO)

**DOI:** 10.1101/2022.12.15.22283543

**Authors:** José Manuel García Almeida, Diego Bellido Guerrero, Daniel de Luis Román, Germán Guzmán Rolo, Gabriel Olveira

**Affiliations:** Department of Endocrinology and Nutrition, Hospital Universitario Virgen de la Victoria, IBIMA, CI-BEROBN, Hospital Quirónsalud, University of Málaga, Spain; Department of Endocrinology and Nutrition, Complejo Hospitalario de Ferrol, A Coruña, Spain; Department of Endocrinology and Nutrition, Institute of Endocrinology and Nutrition, Medicine School and Department of Endocrinology and Investigation, Hospital Clínico Universitario, University of Valladolid, Valladolid, Spain; Medical department. Abbott Laboratories. Spain; Department of Endocrinology and Nutrition. Regional University Hospital of Málaga. University of Málaga. IBIMA BIONAND Platform of Málaga, Spain

**Keywords:** nutritional ultrasound, nutritional biomarker, ultrasound cut-off values, disease-related malnutrition, GLIM, SGA, body composition, sarcopenia, quadriceps femoris muscle, abdominal muscle area, muscle mass

## Abstract

**Background:** The application of nutritional ultrasound for the morphological and structural study of muscle mass is an emerging technique in clinical nutrition. Currently, all definitions of malnutrition include the measurement of muscle mass involvement, however, there is no single way to assess it. It is necessary to develop new techniques to identify muscle involvement in malnutrition that are valid, standardized, reliable, accurate and profitable.

**Objectives:** The objective of this study is to value the new muscle ultrasound techniques aimed to measure muscle and functional status, to make a more accurate diagnosis and a better prediction of complications and morbidity and mortality in patients at nutritional risk. Primary outcome: to assess the feasibility of ultrasound or muscle ultrasound techniques in both nutritional diagnosis and follow-up, over 3 to 6 months, in a nutritional intervention program.

**Methods:** DRECO (Disease-Related caloric-protein malnutrition EChOgraphy) is a prospective, multicenter, uncontrolled clinical study in standard clinical practice to value the usefulness of nutritional ultrasound (muscle ultrasound) in the nutritional diagnosis and follow-up of patients over a period of 3 to 6 consecutive months, after standard nutritional clinical practice intervention, and physical activity to control their disease-related malnutrition.

**Discussion:** This study will standardize nutritional ultrasound measures. It will validate and define specific cut-off values for nutritional ultrasound and get its correlation with already well-known nutritional tools such as SGA (Subjective Global Assessment) or GLIM (Global Leadership Initiative on Malnutrition) criteria. Thus, muscle ultrasound will become not only a tool to assess the diagnosis of malnutrition, but it will be integrated in the routine clinical practices to evaluate nutritional interventions.

**Registration:** This study is registered at clinicaltrials.gov (NCT05433831), registered on June 27^th^, 2022. https://clinicaltrials.gov/ct2/show/NCT05433831.

## 1. INTRODUCTION

Disease-related malnutrition (DRM) can occur when there is a deficient supply of energy, protein and/or other nutrients, depending on the nutritional needs of everyone at different times of their life cycle or health or disease circumstances. This deficiency induces effects on body composition and tissue and organ function and results in clinical consequences: increased morbidity and mortality associated with different disease processes (1).

In 2019, the GLIM criteria were published (2), providing a different vision of how to assess the malnourished patient. These criteria are divided into both phenotypic and etiological criterion:

- Phenotypic criterion
  - Weight loss (%): >5% within past 6 months, or >10% beyond 6 months
  - Low body mass index (kg/m^2^): <20 if < 70 years, or <22 if >70 years. Asia: <18.5 if < 70 years, or <20 if >70 years
  - Reduced muscle mass: Reduced by validated body composition measuring techniques
- Etiological criterion
  - 50% of ER (energy requirements) > 1 week, or any reduction for >2 weeks, or any chronic GI (gastrointestinal) condition that adversely impacts food assimilation or absorption
  - Inflammation: Acute disease/injury or chronic disease-related

There are techniques for nutritional assessment using assessment tools aimed at morpho functional diagnosis of malnutrition (3), in addition to the classical nutritional parameters, such as weight loss, BMI (body mass index), folds, circumferences, albumin, lymphocytes, cholesterol and intake. New advanced parameters are being incorporated into clinical nutrition and their incorporation into clinical practice is of increasing interest, such as measures derived from bioelectrical impedance (BIA) and phase angle (PhA), dynamometry, functional tests, CRP/prealbumin ratio and muscle ultrasound (see Figure 1).

**Figure 1.**
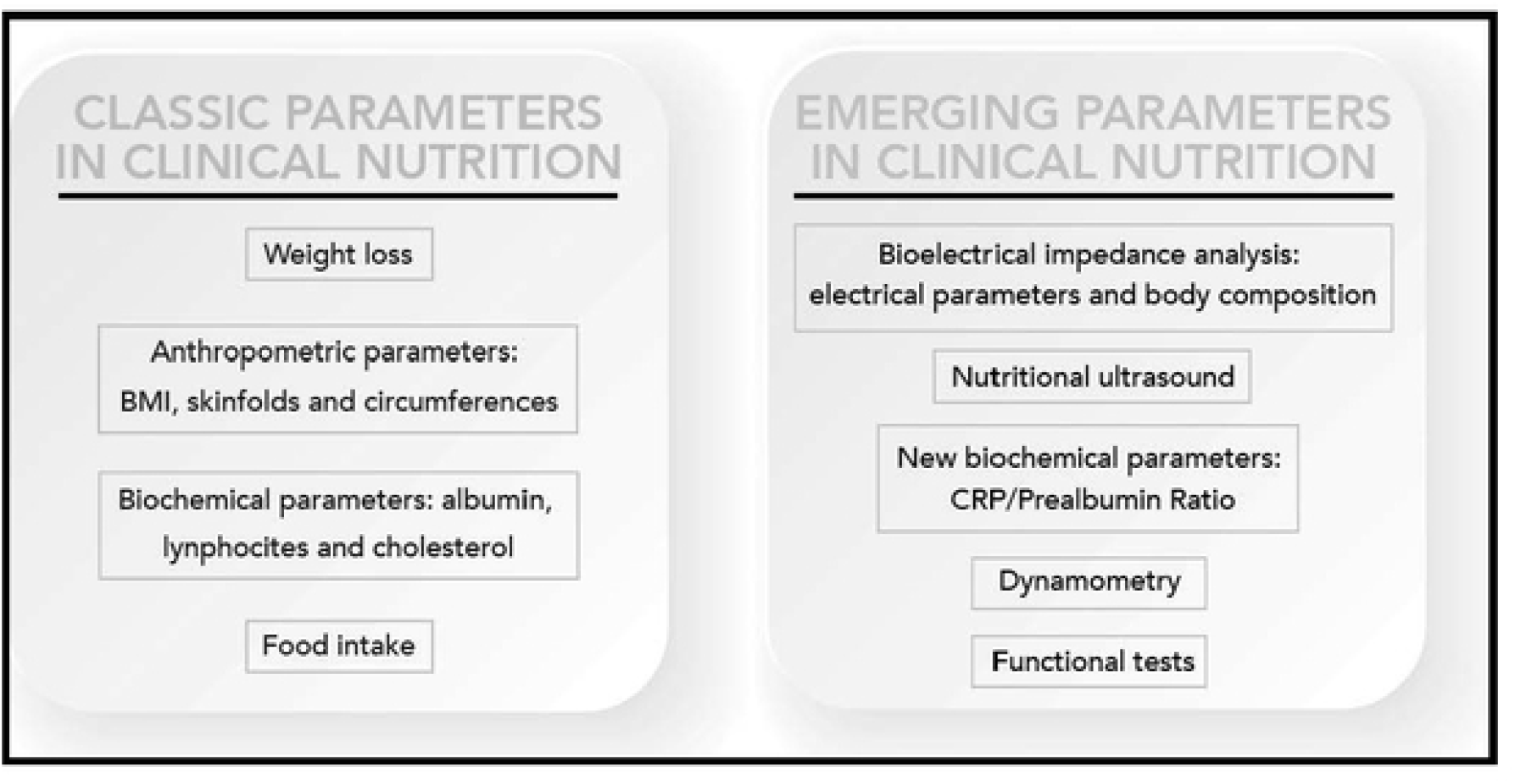
Update of nutritional evolution parameters. Reproduced with permission from the authors (3).

From a scientific point of view, the following nutritional assessment techniques are being incorporated:

### Muscle ultrasound

The application of ultrasound for the morphological and structural study of muscle mass is an emerging technique. Currently, there are different validation studies on the measurement technique. The ultrasound technique determines the surface area of the muscle in transverse and longitudinal position. With ultrasound analysis, it is possible to measure key parameters of muscle architecture, such as muscle volume and muscle fascicle length. Although there are different muscle structures that can be evaluated, many of the studies focus on the quadriceps rectus femoris or on combinations of various muscle groups involving large muscle bundles with functional importance to the patient in terms of gait. Measurement of the rectus femoris of the quadriceps is one of the most referenced measurements due to its correlation with strength and tests of execution or functional performance. It is necessary to develop new techniques to identify muscle involvement in malnutrition that are valid, standardised, reliable, accurate and profitable. Currently, all definitions of malnutrition include the measurement of muscle mass involvement, however, there is no single way to assess it. The classic imaging techniques such as DEXA (dual-energy x-ray absorptiometry), CT (computerised tomography) and MRI (magnetic resonance imaging) are considered the gold standard, but they have difficulties in their clinical application under normal practice conditions. Ultrasound has the advantage of being inexpensive, portable, and does not involve ionising radiation. Several studies have confirmed the reliability of this technique to measure the size of the quadriceps muscle in a healthy population (4). Studies on the reliability of rectus femoris ultrasound have been published with an intraclass coefficient of variation (ICC) of 0.97 (95% CI: 0.92-0.99) for the test-retest reliability of ultrasound.

The American Society for Parenteral and Enteral Nutrition (ASPEN), among the criteria for the diagnosis of malnutrition in adults, recommends including an evaluation of fat and muscle deposits. Specialists must incorporate techniques that properly help to identify the loss of muscle and fat mass for a correct diagnosis of malnutrition. Implementing these evaluation techniques and instruments is challenging and remains a work in progress (5). Muscle ultrasonography correlates with body composition measurement techniques such as BIA and anthropometry in patients with cancer (6). In adults with cystic fibrosis muscle ultrasound measurements, particularly the mean muscular area rectus anterior (MARA), are related to the nutritional status and respiratory function of these patients. (7) The Global Leadership Initiative on Malnutrition (GLIM) has recently appointed a working group to provide consensus-based guidance on assessment of skeletal muscle mass and its role in the malnutrition diagnostic and assessment process. They support the use of US (ultrasound), particularly in settings where its practical applicability provides potential for patient follow-up through repeated measurements, but it requires standardisation through experienced operators, and repeated measurements performed by the same individual. They also encourage further validation studies for the US (8).

### Bioelectrical impedance (BIA)

BIA is used as a tool to obtain data that helps to better understand the patient’s nutritional status, being a non-invasive, inexpensive, and easily transportable technique. Vector analysis and phase angle provide direct data, not being necessary to be later adjusted using formulas or mathematical models, as it is needed with simple or multifrequency bioelectrical impedance or multifrequency (9). This method is based on the analysis of the two bioimpedance vectors: resistance (R) and capacitive reactance (Xc). Resistance is defined as the opposition to a flow of electric current through a circuit component, medium, or substance, providing information about biological fluids, and therefore, related to tissue hydration. A decrease in the resistance/height ratio will indicate swelling or third space; conversely, an increased ratio will indicate dehydration. Reactance is the effect on an electrical current caused by a material’s ability to store energy in cell membranes, so it is related to the cell mass and the integrity of its membranes. A decrease in Xc indicates loss of cell mass. This cell mass is the sum of all metabolically active cells, being the central parameter in the evaluation of nutritional status since the reduction of cell mass is typically related to malnutrition (10).

A recent study conducted by Fernandez-Jimenez, *et al* found that a low SPhA (standardised phase angle)-malnutrition value (SPhA < −0.3) was significantly associated with a higher mortality hazards ratio (HR 7.87, 95% CI 2.56–24.24, p < 0.001). This biological marker could therefore be incorporated among the screening tools and mortality risk assessment in this population (11).

### Dynamometry

Dynamometry is one of the 6 criterions to define malnutrition according to ASPEN (12). It is extremely sensitive to nutritional status changes, so it is particularly useful to track nutritional therapy or interventions results, even in the short and medium term. It has mostly been used to predict post-surgical complications including elderly patients (13). Results obtained are compared to the population averages by age and sex. Sanchez et al (14) presented reference values for hand dynamometry using a Jamar hand dynamometer for a Spanish population, providing cut-off points to define malnutrition. They concluded that hand dynamometry is associated with lean mass, which supports its usefulness in nutritional assessment.

Although the new GLIM consensus-based guidance on assessment of skeletal muscle mass do not include dynamometry as a marker of muscle mass (8), the authors hereby signing this article have previously studied dynamometry as a marker of muscle mass suggesting that GLIM criterion and dynamometry are associated to a higher mortality rate in both hospitalised and outpatient oncology patients (15, 16).

### Functional tests

These tests are a series of physical activities related to mobility, walking or balance. Their results are related to those of scales that assess instrumental activities of daily living (IADL). The most common are the “Timed Up and Go test” (TUG), the “Gait Speed Test” (GST) and the “Short Physical Performance Battery (SPPB)” test that includes 3 tests (balance, gait speed and get up and walk) (17).

Besides, the decrease in physical performance, evaluated by the SPPB test or hand grip strength, has been shown to be elevated in patients with colorectal cancer prior to surgery and it was related to an increase in postoperative complications and mortality (18).

### 1.1. STUDY OBJECTIVES

The objective of this study is to value the new muscle ultrasound techniques aimed to measure muscle and functional status, to make a more accurate diagnosis and a better prediction of complications and morbidity and mortality in patients at nutritional risk. This main objective is developed in primary and secondary objectives as it follows:

#### 1.1.1. Primary objective

- To assess the feasibility of ultrasound or muscle ultrasound techniques in both nutritional diagnosis and follow-up, over 3 to 6 months, in a nutritional intervention programme.

#### 1.1.2. Secondary objectives

- To determine the association between muscle morphological parameters (nutritional ultrasound of the leg (area, circumference, axis and adipose tissue), total abdominal and pre-peritoneal parameters measured by nutritional ultrasound and the nutritional and functional status of the patient, as well as their prognostic value in hospitalised patients.
- To establish an association between ultrasound as a diagnostic value of malnutrition as compared to the diagnostic gold standard (SGA and GLIM criteria).
- To determine the ultrasound cut-off points associated with the diagnosis of malnutrition and sarcopenia using the following tools:
  - Measurement of body composition using impedance techniques (Report: Phase angle, body cell mass (BCM), hydration, fat free mass (FFM) and lean mass index.
  - Muscle strength and capacity to perform physical activity after the intervention: dynamometry and Timed Up and Go test (TUG).
  - Criteria for sarcopenia.
  - To assess association with inflammatory activity markers: High-sensitivity C-reactive protein (CRP)/prealbumin.
- To assess ultrasound changes in patient follow-up.
- To establish an association of ultrasound results as predictors of morbidity and mortality (stay, mortality at 3 and 6 months, readmissions and in-hospital complications).

## 2. PATIENT PARTICIPANT INVOLVEMENT AND FEASIBILITY OF STUDY DESIGN

DRECO (Disease-Related caloric-protein malnutrition EChOgraphy) is a prospective, multicentre, uncontrolled clinical study in standard clinical practice to value the usefulness of nutritional ultrasound (muscle ultrasound) in the nutritional diagnosis and follow-up of patients over a period of 3 to 6 consecutive months, after standard nutritional clinical practice intervention, and physical activity to control their disease-related malnutrition.

The study may be considered non-interventional since patients will undergo nutritional interventions and the standard treatment planned by their physician for treatment according to his/her standard clinical practice, and the only addition to the standard measurement and follow-up techniques of the patient will be the performance of a muscle ultrasound measurement using equipment provided to the centre for this purpose.

Patients over 18 years of age who, in the first week of hospital admission in medical-surgical areas, excluding critical patients, have an assessment of risk of malnutrition according to the MUST and SARC-F screening test using R-MAPP. [(MUST: Malnutrition Universal Screening Tool; SARC-F is an acronym of 5 domains included in the questionnaire: 1) Strength, 2) Assistance with walking, 3) Rising from a chair, 4) Climbing stairs, and 5) Falls; R-MAPP (Remote consultation on MAlnutrition in the Primary Practice)].

If the results show a moderate or high risk of malnutrition, these patients will be invited to participate in the study, and will undergo the morpho functional assessment, an ultrasound study and the Subjective Global Assessment (SGA). This study is registered under ClinicalTrials.gov (NCT05433831).

Figure 2 shows the schedule of enrolment, interventions, and assessments.

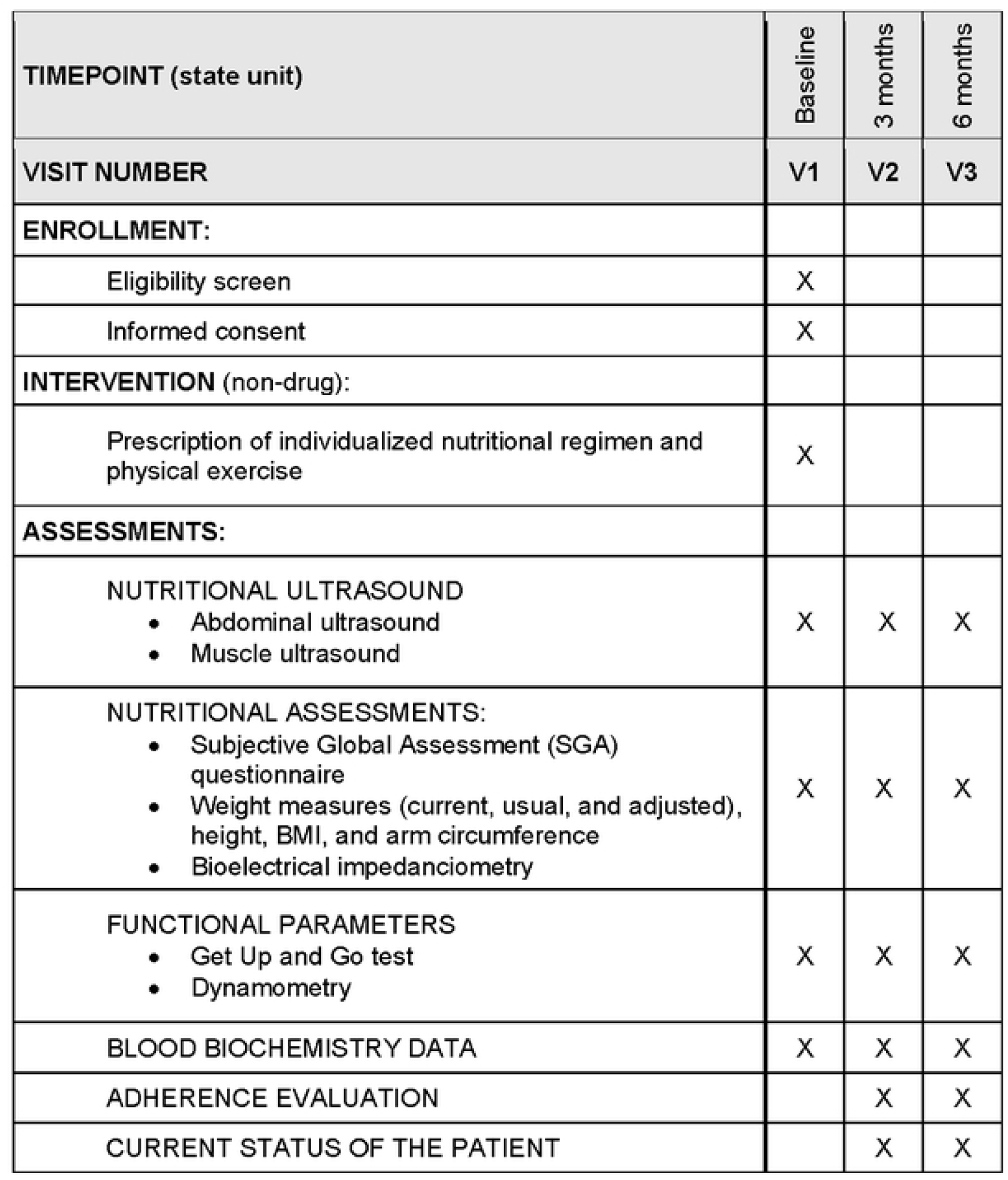

### 2.1. Inclusion criteria

- Patients admitted to hospital who in the first week of admission have moderate or high risk of malnutrition according to the MUST and SARC-F screening test using R-MAPP.
- Patients aged 18 to 85 years.
- Patient who agrees to participate in the study and signs the informed consent.

### 2.2. Exclusion criteria

- Hepatic impairment - AST/ALT (aspartate aminotransferase/alanine aminotransferase) 3 x upper limit of normal.
- Chronic kidney failure - GFR (glomerular filtration rate) <45 mL/min).
- Patients with previous ICU (intensive care unit) stay during the study admission.
- Cancer patients on palliative treatment or ECOG (Eastern Cooperative Oncology Group) ≥ 3.
- Orthopaedic disease that does not allow adequate walking.
- Patients with known dementia or others not related to a significant neurological or psychiatric disorder, or any other psychological condition that may interfere with the conduct of the study.
- Patients with eating disorders.
- Life expectancy of less than 6 months.
- Patients unable to adequately complete the clinical laboratory assessments required for the study protocol.

### 2.3. Sample size calculation

There are no previous clinical trials focusing on this objective published in the literature. We report a study in patients with chronic kidney disease on haemodialysis (HD) (19) where measurement of the rectus femoris cross-sectional muscle area (RFCSA) was validated for the diagnosis of malnutrition related to this condition. RFCSA compared to bioimpedance spectroscopy had higher area under the curve (AUC, 0.686 vs. 0.581), sensitivity (72.8% vs. 65.8%), and specificity (55.6% vs. 53.9%). The AUC of RFCSA was higher for the risk of protein-energy wasting (PEW) in male (0.74, 95% CI: 0.66 to 0.82) and female patients (0.80, 95% CI: 0.70 to 0.90) (both p<0.001). Gender-specific RFCSA values (males <6.00 cm2; females <4.47 cm^2^) indicated that HD patients with lower RFCSA were 8 times more likely to have PEW (AOR = 8.63, 95% CI: 4.80-15.50, p<0.001).

Our study aims to establish the feasibility of nutritional ultrasound measurements at different ages in both sexes to apply to patients with nutritional risk worldwide. For this purpose, the electronic CRF will be programmed with the sample distributed by quotas to cover 50% men and 50% women, as well as 10-year age ranges. Age-stratified sampling is designed to obtain representative results of different ages and could be associated with the results of VGS, BIA, and dynamometry. Variability of measurements should be adjusted for sex, age and anthropometric parameters such as height.

It is estimated that 1,000 patients with nutritional risk will be discharged from 20-25 healthcare centres throughout Spain and that at least 60% of the population will complete the 3-to-6-month follow-up of the study. Due to the special pandemic situation, a higher-than-expected drop-out rate is expected at 6 months than under normal conditions (40% are estimated not to complete the 6-month follow-up for any reason).

### 2.4. Study conduct

The physicians participating in the study will be responsible for assessing the suitability of inclusion for each patient.

Patients will be consecutively recruited by the physician as they are assessed daily in their clinical practice at the hospital and found to have a risk of malnutrition according to the MUST/SARC-F (R-MAPP) screening test.

Before inclusion, the investigator must check the inclusion and exclusion criteria and obtain their informed consent.

The physician will be responsible for applying nutritional intervention and physical activity treatment according to standard clinical practice, as well as for clinical monitoring of patients. The treatment prescribed to each patient is not the objective of this study and is how the patient will experience changes that must be recorded with the different techniques described and with the muscle ultrasound involved in this study.

All physicians participating in the study must have been previously trained in the use of the ultrasound equipment and materials provided for the study, as well as in the use of the electronic CRF for data entry designed for this study.

#### 2.4.1. Nutritional ultrasound techniques and measurements

(ultrasound with 4-10 cm linear tube). The equipment provided for the study is UProbe L6C Ultrasound Scanner (linear transducer 7.5-10 kHz) that allows depths up to 100 mm. Manufactured by Guangzhou Sonostar Technologies Co., Ltd. PR China.

##### 2.4.1a. Quadriceps rectus femoris ultrasound (see Figure 3)

The measurement technique is determined for the patient lying supine with knees extended and relaxed.

**Figure 3.**
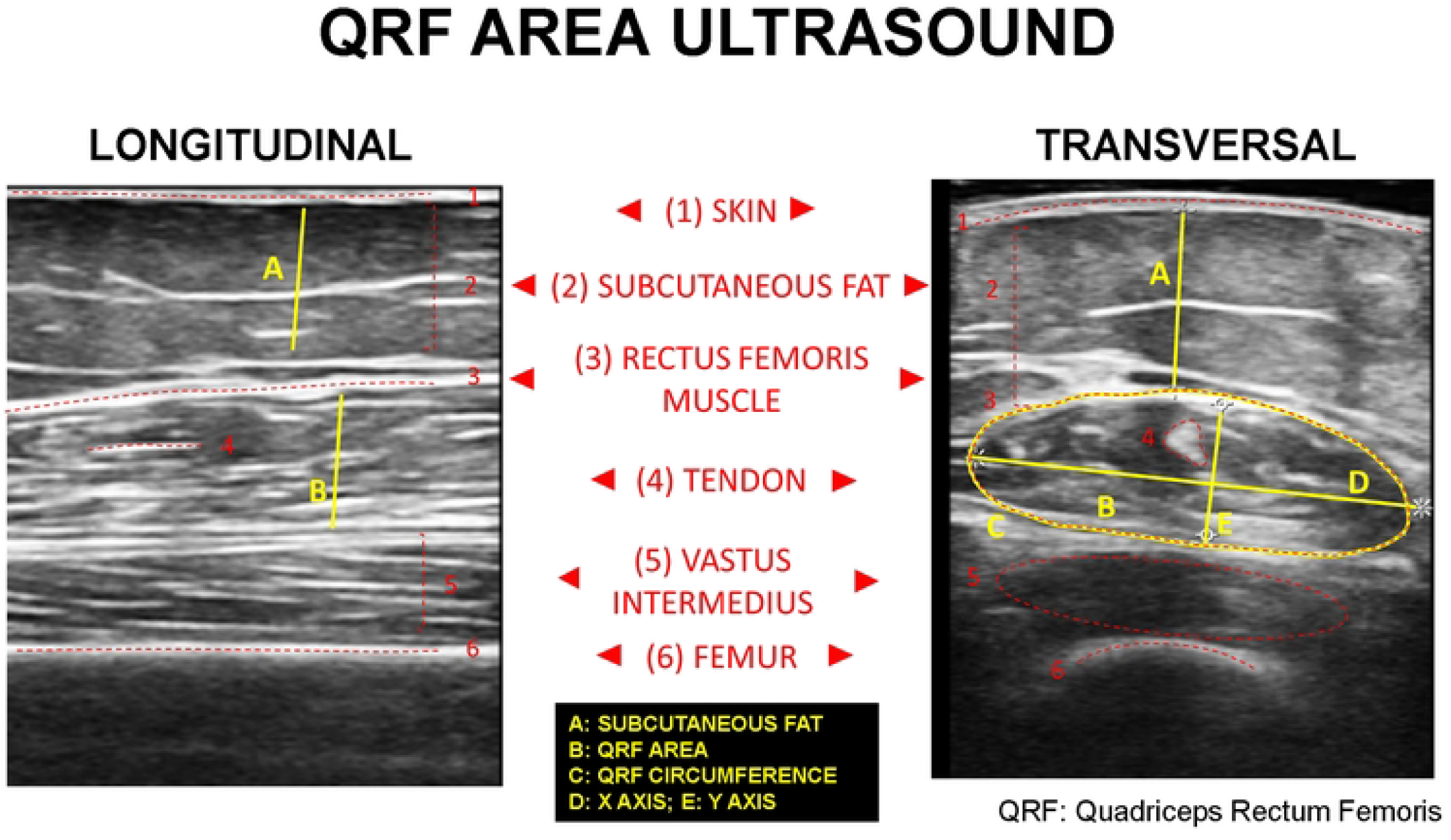
Comparison of longitudinal and transversal sections of the QRF muscle area ultrasound. Functional measures and main anatomical structures are represented.

###### Measurement technique

- In the lower third of the imaginary line between the antero-superior iliac spine and the superior border of the patella.
- Correction of leg angle, it is important to focus the image on the rectus femoris.
- In malnourished patients, loss of muscle tone causes the muscle to move to the sides
- To minimize variability, measurements must be repeated three times.

##### 2.4.1b. Abdominal ultrasound (see Figure 4)

Total, superficial, and pre-peritoneal adipose tissue are measured (centimetres) for the patient lying down.

**Figure 4.**
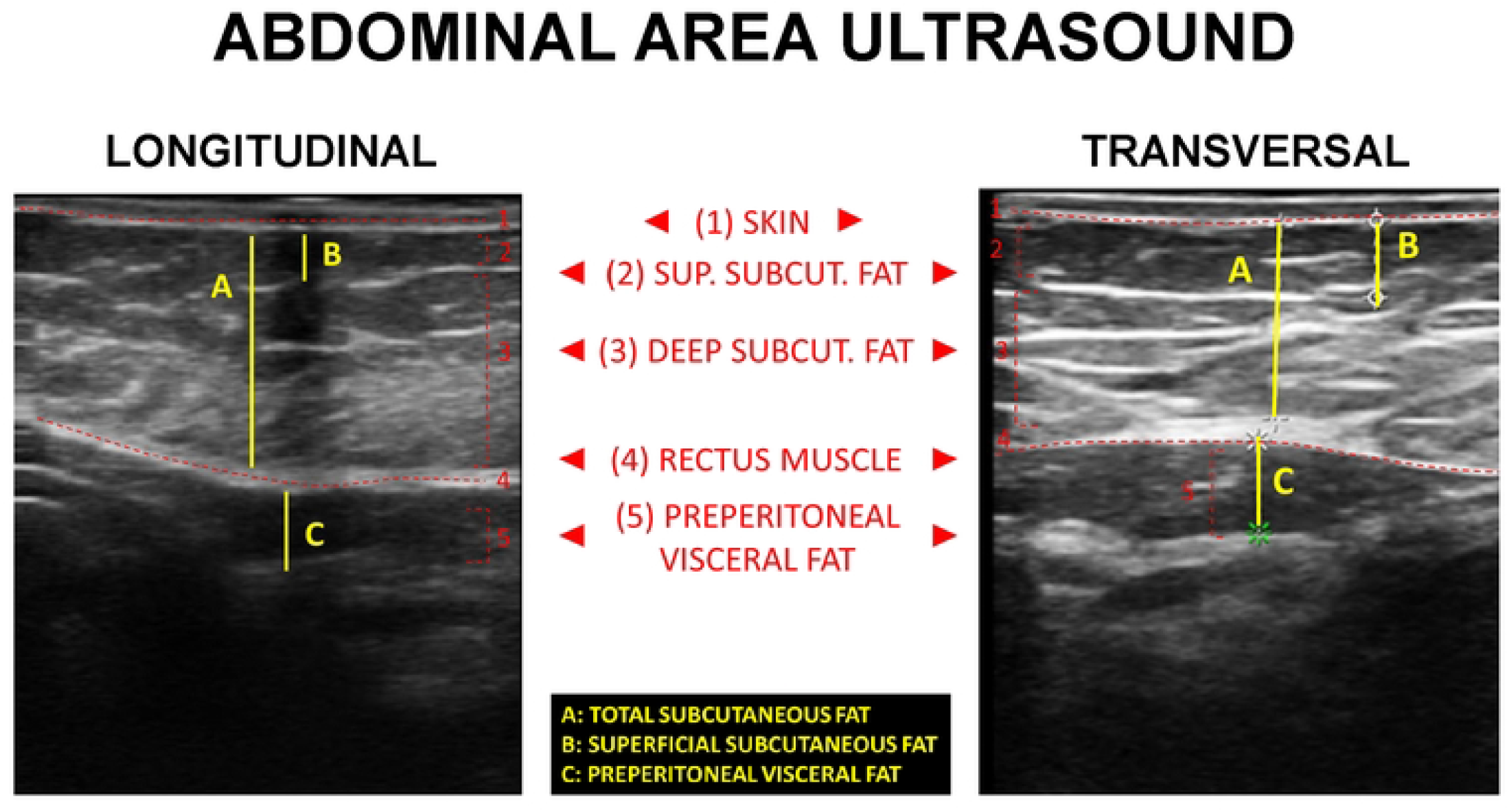
Comparison of longitudinal and transversal sections of the QRF muscle area ultrasound. Functional measures and main anatomical structures are represented.

###### Measurement technique

- The transducer is placed between the xiphoid process and the umbilicus in the midline (in patients with surgery without navel, this would be 10 cm from the xiphoid appendix).
- Images are taken during non-forced expiration, in a transverse plane with a variable probe depth of 4-10 cm, perpendicular to the skin.
- To minimize variability, measurements must be repeated three times.

###### Measurement planes

- Measurement of subcutaneous adipose tissue: the superficial and deep layers are differentiated.
- Visceral adipose tissue measurement: it is measured in a transverse position. Measure the distance between the boundary of the parietal peritoneum to the *linea* alba on the inner side at the junction of the two-rectus straight abdominis muscles.

#### 2.4.2 Follow-up period

The planned follow-up period for each patient will be 3 to 6 months from the inclusion visit.

The investigating physician will perform at least one first inclusion visit, and a follow-up visit at 3 and 6 months for each patient.

#### 2.4.3. Study duration

The study is planned to last 18 months to detect patients at risk of malnutrition, recruitment, field work, monitoring and data analysis.

An estimated 2-3 months will be needed to plan the coordination and distribution of the work in the hospitalisation and outpatient clinic areas for the selection of candidate patients. It will take 6 to 9 months to recruit patients. From the start of the study, the database will be completed, and preliminary analyses will be performed. The final analysis will be performed when the follow-up is completed together with writing of the related work that will require 4 to 6 months to complete.

### 2.5. Outcome measures

A list of the outcomes of interest is provided in Table 1.

| PRIMARY OUTCOMES |  |
| --- | --- |
| Nutritional ultrasound measurements: ultrasound with 4-10 cm linear probe * |  |
|  | Abdominal ultrasound: total, superficial and pre-peritoneal adipose tissue (measured in centimetres) |
|  | Muscle ultrasound: Area, circumference, axes and adipose tissue (measured in centimetres) |
| SECONDARY OUTCOMES |  |
| Sociodemographic data: |  |
|  | Age<br>Sex<br>Educational level<br>Toxic habits<br>Medical history<br>Risk of sarcopenia and moderate to high malnutrition based on MUST and SARC-F screening test using R-MAPP |
| Subjective Global Assessment (SGA) questionnaire |  |
| Anthropometric data: |  |
|  | Current body weight (measured or estimated)<br>Usual weight<br>Adjusted weight (adjusted weight in obese subjects, dry weight without oedema in malnourished subjects)<br>Height (measured or estimated)<br>BMI<br>Arm circumference |
| Bioelectrical impedance data (model (50 kHz): ** |  |
|  | TBW (total body water, L)<br>ECW (extracellular water, L)<br>ICW (intracellular water, L)<br>FFM (lean mass, kg)<br>FM (fat mass, kg)<br>BCM (body cell mass, kg)<br>ASMM (appendicular skeletal muscle mass, kg)<br>SMI (skeletal muscle mass index, kg)<br>Percent hydration<br>Body fat (%) |
| Blood biochemistry data (at baseline visit, at 3 and 6 months): |  |
|  | Albumin<br>Prealbumin<br>C-reactive protein<br>Total cholesterol<br>Lymphocytes |
| Bioelectrical impedance data (model (50 kHz): ** |  |
|  | Age<br>Sex<br>Educational level<br>Toxic habits<br>Medical history<br>Risk of sarcopenia and moderate to high malnutrition based on MUST and SARC-F screening test using R-MAPP |
| Functional parameters |  |
|  | Timed Up and Go test (TUG): patient sits in a chair and is told to get up (timing starts), walks 3 metres, comes back and sits in the initial chair (timing ends). Interpretation: <20 seconds: normal, > 20 seconds: increased risk of falling.<br>Dynamometry. Three measurements of the dominant hand will be made recording the mean and maximum, measured in kilograms. Jamar® dynamometers are most used in international studies and have several grip positions. |
| Current patient status |  |
|  | Hospital stays, mortality at 3 and 6 months, hospital readmissions and complications, if occurring, and their consequences (resolved/unresolved) must be recorded in the form. |
| Adherence |  |
|  | Attendance to study follow-up visits. |

\*The equipment provided for the study is the UProbe L6C Ultrasound Scanner (linear transducer 7.5-10 kHz) that allows depths up to 100 mm. Manufactured by Guangzhou Sonostar Technologies Co., Ltd. PR China.
\*\*Each healthcare center could use the BIA device they already owned. The most used device among all participants was AKERN branded.

### 2.6. Data analysis plan

Data analysis will be performed using SPSS 22.0 software (SPSS Inc., Chicago, IL, USA).

Quantitative variables will be expressed as mean ± standard deviation. The comparison between qualitative variables will be performed using the Chi-square test with Fisher’s correction when necessary. Quantitative variables will be analysed using a Kolmogorov-Smirnov test. Differences between quantitative variables will be analysed using Student’s t or ANOVA tests (for two or more samples, respectively) and non-parametric tests (Mann-Whitney or Kruskal-Wallis) will be used when the variables to be analysed do not follow a normal distribution.

Kappa coefficient will be used to assess agreement between techniques in diagnosis of malnutrition.

The association between variables will be studied using Spearman or Pearson correlations according to normality.

Several cut-off points will be estimated for prediction of diagnosis of malnutrition and sarcopenia using ultrasound by ROC curves.

The significant associations between muscle ultrasound parameters and the objective clinical variables in the univariate analysis will then be analysed in multivariate logistic regression models which also control other confounding variables. To assess which nutritional tool best predicts the risk of mortality during admission (and re-admission), we will perform multivariate logistic regression models, in which the dependent variable will be in-hospital mortality (or re-admission) based on the different tools applied (e.g. ultrasound, phase angle, SGA criteria, GLIM, LMI), also controlling for sex, the presence of previous comorbidities and other variables showing association in the univariate study.

For all calculations, a probability p less than 0.05 for two tails will be considered significant.

#### 2.6.1. Recording of adverse reactions

Adverse reactions reporting is not the objective of the study. The investigator should proceed as usual and through the channels established in the healthcare system if any adverse effect occurs during follow-up. It will only be recorded in the follow-up if the patient must leave the study for this reason for statistical purposes.

#### 2.6.2. Handling of missing data

No formal imputation will be made for the different analyses; therefore, all estimates will be obtained using all available data (available data only, ADO).

Since the study will be recorded using an electronic CRF (case report form), the necessary consistency filters and alerts for missing data will be programmed to validate and store the information, in order to minimise missing data and prevent the entry of incorrect or out of range data.

## 3. ETHICS

### 3.1. General aspects

This study will be conducted in accordance with current regulations, accepted international ethical standards of Good Clinical Practice (CPMP/ICH/135/95), the principles laid down in the latest version of the Declaration of Helsinki, RD 1591/2009 and Circular No. 07/2004 regulating clinical research with medical devices.

### 3.2. Informed consent

Before inclusion in the study and after considering the suitability of patient inclusion, all participating physicians must offer the patient information about the study using a patient information sheet, invite the patient to participate in it, answer their questions and request completion of the informed consent form that will be kept in their own file.

### 3.3. Evaluation by an Ethics Committee

All study materials will be submitted to the Provincial mpREC of Málaga for evaluation (Area of the Regional University Hospitals of Malaga and Virgen de la Victoria). The study will be initiated after IRB/IEC approval is obtained.

Provincial mpREC of Malaga.

Regional University Hospital of Málaga

Wing A. General Hospital, 7th floor 9010, Málaga.

### 3.4. Confidentiality

The study data will be entered into an automated file owned by the sponsor. The analysis of study results will be made from an anonymised database, that is, dissociated, with no personal data, so that no subject can be identified or identifiable. This study database will be extracted from the electronic CRF and will include data from physician records, impedance recordings, and muscle ultrasound images. Data from different sources will be linked from the patient code and will not include personal data. All data in the file owned by the sponsor will be treated confidentially. The sponsor undertakes not to transfer data to third parties.

## 4. DISCUSSION

There is a growing interest in the literature on the evaluation of muscle mass by ultrasound (20). Its current clinical utility focuses on measuring the involvement of muscle mass to assess the nutritional status of a patient (21). The further step that it is being investigated in this clinical study, is that muscle ultrasound becomes not only a tool to assess the diagnosis of malnutrition but to integrate it in the routine clinical practices to evaluate nutritional interventions.

The evaluation of the nutritional ultrasound should enable clinical decisions based on its results to permit the adjustment and individualization of the nutritional therapeutic and physical exercise plan, along with functional recovery (20).

To the best of our knowledge, this is going to be the largest study (sample size=1,000) using nutritional ultrasound in patients with nutritional risk. Current scientific evidence is limited, and it is expected that such a large population will allow us to validate and define specific cut-off values for nutritional ultrasound and get its correlation with already well-known nutritional tools such as SGA or GLIM criteria (22).

The emerging field of ultrasound assessment of muscle mass only highlights the need for a standardisation of measurement technique as Perkisas, *et al* outline in their recently published 2022 SARCUS update. This update provides the approach of muscle assessment according to the most recent literature and anatomical landmarks for 39 different muscles. Besides, the discussion about 4 new muscle parameters that are added to the 5 that were previously considered is also presented (23) and some of these parameters have been correlated with PhA (24) and they will be analysed in our present protocol. Our ongoing study is intended to standardize these outstanding technique measures, to apply this technique widely soon.

## Data Availability

No datasets were generated or analysed during the current study. All relevant data from this study will be made available upon study completion.

## Author Contributions

All authors have identified the research question and were responsible for the conception and design of the protocol and the study. JM.G.A., D.B.G, D.L.R, and G.O.F. are conducting study investigation. G.G.R has managed funding acquisition. All authors have been involved in drafting the manuscript and revising it critically for intellectual content. All authors read and approved the final manuscript.

## Institutional Review Board Statement

The study was conducted in accordance with the Declaration of Helsinki, and approved by the following Ethical Committees: Comité de Ética de la In-vestigación Provincial De Málaga, Comité de Ética de la Investigación con medicamentos Área De Salud Valladolid, Comité de Ética de la Investigación provincial de Córdoba, Comité de Ética de la Investigación con medicamentos del Complejo Hospitalario Universitario de Canarias (Provincia de Santa Cruz de Tenerife), Comité de Ética de la Investigación con medicamentos del Complejo Hospitalario Universitario de Las Palmas, Comité de Ética de la Investigación con medicamentos y Comisión de Proyectos de Investigación del Hospital Universitari Vall D’Hebron, Comité de Ética de la Investigación con medicamentos del Consorcio Hospital General Universitario de Valencia, Comité de Ética de la Investigación con medicamentos de la Clínica de Navarra, Comité de Ética de la Investigación con medicamentos de Euskadi and Comité de Ética de la Investigación con medicamentos del Hospital Universitario y Politécnico la Fe de Valencia. This study is registered at clinicaltrials.gov (NCT05433831), registered on June 27th, 2022. https://clinicaltrials.gov/ct2/show/NCT05433831.

## Informed Consent Statement

All participants are provided with a participant information sheet and are required to provide written consent.

